# Functional characterization of six *SLCO1B1* (OATP1B1) variants observed in Finnish individuals with a psychotic disorder

**DOI:** 10.1101/2022.08.10.22278615

**Authors:** Katja Häkkinen, Wilma Kiander, Heidi Kidron, Markku Lähteenvuo, Lea Urpa, Jonne Lintunen, Kati-Sisko Vellonen, Seppo Auriola, Minna Holm, Kaisla Lahdensuo, Olli Kampman, Erkki Isometsä, Tuula Kieseppä, Jouko Lönnqvist, Jaana Suvisaari, Jarmo Hietala, Jari Tiihonen, Aarno Palotie, Ari V. Ahola-Olli, Mikko Niemi, the SUPER-Finland researchers listed in the Acknowledgements

## Abstract

**Aim:** Variants in the *SLCO1B1* (solute carrier organic anion transporter family member 1B1) gene encoding the OATP1B1 (organic anion transporting polypeptide 1B1) protein are associated with altered transporter function that can predispose patients to adverse drug effects with statin treatment. We explored the effect of six rare *SLCO1B1* single nucleotide variants (SNVs) occurring in Finnish individuals with a psychotic disorder on expression and functionality of the OATP1B1 protein.

**Methods:** The SUPER-Finland study has performed exome sequencing on 9,381 individuals with at least one psychotic episode during their lifetime. *SLCO1B1* SNVs were annotated with PHRED-scaled Combined Annotation-Dependent (CADD) scores and the Ensembl variant effect predictor (VEP). *In vitro* functionality studies were conducted for the SNVs with PHRED-scaled CADD score >10 and predicted to be missense. To estimate possible changes in transport activity caused by the variants, transport of 2’,7’-dichlorofluorescein (DCF) in OATP1B1 expressing HEK293 cells was measured. The amount of OATP1B1 in crude membrane fractions was quantified with a LC–MS/MS-based quantitative targeted absolute proteomics analysis.

**Results:** Six rare missense variants of *SLCO1B1* were identified in the study population located in transmembrane helix 3: c.317T>C (p.106I>T), intracellular loop 2: c.629G>T (p.210G>V), c.633A>G (p.211I>M), c.639T>A (p.213N>L), transmembrane helix 6: 820A>G (p.274I>V), and the C-terminal end: 2005A>C (p.669N>H). Of these variants, *SLCO1B1* c.629G>T (p.210G>V) resulted in protein loss of function, abolishing the uptake of DCF and reducing membrane protein expression to 31% of reference OATP1B1.

**Conclusions:** Of the six rare missense variants, *SLCO1B1* c.629G>T (p.210G>V) causes loss-of-function of OATP1B1 transport and severely decreases membrane protein abundance. Carriers of *SLCO1B1* c.629G>T may be susceptible to altered pharmacokinetics of OATP1B1 substrate drugs and may have increased likelihood of adverse drug effects such as statin-associated musculoskeletal symptoms.

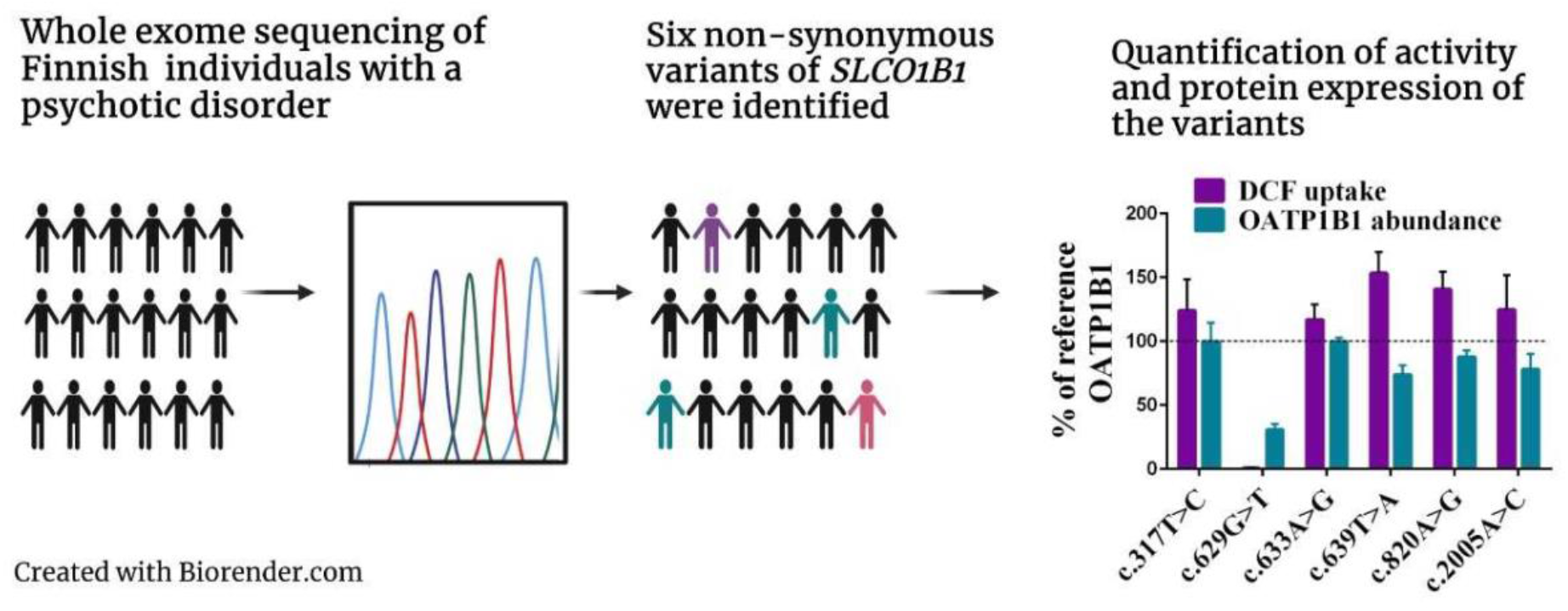

## Introduction

Statins are among the most common prescription drugs in the world and are used for the reduction of low-density lipoprotein cholesterol concentration and prevention of cardiovascular disease. The solute carrier organic anion transporter family member 1B1 (*SLCO1B1)* gene encodes the organic anion transporting polypeptide 1B1 (OATP1B1), a transmembrane protein that is involved in the transport of drugs such as statins and other compounds from the blood into the hepatocytes^1,2^.

Single nucleotide variants (SNVs) in *SLCO1B1* are associated with impaired transporter function that can alter systemic exposure to statins. This can consequently predispose patients to adverse drug effects including statin-associated musculoskeletal symptoms (SAMS) and even rhabdomyolysis, impacting statin adherence and hindering the long-term effectiveness of statin therapy^3,4,5,6,7^. Cardiovascular mortality is one of the leading causes of excess mortality in patients with schizophrenia^8^. Antipsychotic medications, especially second-generation antipsychotic medications such as clozapine and olanzapine, can induce metabolic abnormalities including dyslipidemias^9^. Therefore, statins are commonly prescribed to patients with psychotic disorders.

The U.S. Food and Drug Administration (FDA) and European Medicines Agency (EMA) acknowledge the significance of OATP1B1 variants in medication safety by providing guidelines for studying the drug transporter’s effect on pharmacokinetics in drug development^10,11^.

Pharmacogenetic research has suffered from small sample sizes, and finding a patient group of sufficient size is often challenging when exploring rare genetic variants^12^. Rare variants of *SLCO1B1* are mainly unreported although their effect on drug safety can be as consequential as the effect of common variants^13,14^. Interpretation of genetic variants from vast amounts of whole exome sequencing data remains a challenge and computational tools are needed to predict the functional impact of pharmacogenetic variants before the identified variants can be explored at a biological level^15,16,17^. Various algorithms that are needed to predict the functional impacts of coding variants are available using standard criteria and scoring metrics or machine learning approaches. Biological assessment of *in silico* evaluated variants is necessary to generate clinically actionable recommendations.

We studied the potential clinical relevance of rare *SLCO1B1* SNVs occurring among 9,381 Finnish individuals with a psychotic disorder recruited as part of the SUPER-Finland study. The variants were selected for *in vitro* expression and functionality experiments based on a damaging *in silico* prediction by the PHRED-scaled Combined Annotation-Dependent (CADD) score algorithm^18,19^ and an Ensembl variant effect predictor (VEP)^20^ annotation of missense. Cellular uptake of the substrate 2,7-dichloro- fluorescein (DCF) in OATP1B1-overexpressing HEK293 cells was determined and the amount of OATP1B1 variant protein expressed in HEK293 cells was quantified with an LC–MS/MS based quantitative targeted absolute proteomics (QTAP) analysis. Our results provide functional annotations for *SLCO1B1* c.317T>C (p.I106T), c.629G>T (p.G210V), c.633A>G (p.I211M), c.639T>A (p.N213K), c.820A>G (p.I274V) and c.2005A>C (p.N669H) variants.

## Material and methods

### SUPER-Finland study

The SUPER-Finland study recruited 10,474 participants aged >18 with a severe mental disorder from Finnish in- and outpatient psychiatric care, primary care, housing units and additionally with newspaper advertising between the years 2016-2018. Subjects with a diagnosis of a schizophrenia spectrum disorder (ICD-10 codes F20, F22–29), bipolar disorder (F30, F31) or major depressive disorder with psychotic features (F32.3 and F33.3) were included in the study. Exclusion criteria were inability to give informed consent and age under 18 years. We put special focus on ensuring wide coverage of known Finnish internal population subisolates^21^. The SUPER-Finland study protocol will be described in a separate cohort profile manuscript that is in preparation.

DNA was extracted from participants’ blood samples collected by venipuncture (2x Vacutainer EDTA K2 5/4 ml, BD). A saliva sample (DNA OG-500, Oragene) was collected for DNA extraction when venipuncture was not possible. The samples were frozen (−20 °C) within 60 min of sampling and sent to THL Biobank (Finnish Institute for Health and Welfare) within 3 months for long term storage (−185 °C). PerkinElmer Janus chemagic 360i Pro Workstation with the CMG-1074 kit was used to extract DNA from the EDTA-blood tubes. Extraction of DNA from saliva samples (after incubation +50 °C, o/n) was performed by Chemagen Chemagic MSM I robot with the CMG-1035–1 kit. The DNA samples were genotyped and sequenced at the Broad Institute of MIT and Harvard, Boston Cambridge, Massachusetts, USA.

### Exome sequencing of SUPER-Finland study samples

To discover rare *SLCO1B1* SNVs with possible changes in OATP1B1 protein function, we used exome data from 9,381 SUPER-Finland study participants. Exome sequencing was carried out with the Illumina HiSeq platform using 151 base pair paired-end reads at the Broad Institute of Harvard and MIT. The samples were enriched with the Illumina Nextera capture kit and sequenced until 80% of the target capture was covered at 20x. The Picard sequence processing pipeline was used to process BAM files (http://broadinstitute.github.io/picard/) and the data was mapped to the human genome reference build 38 (GRCh38) using BWA^22^. This procedure followed standard best practice alignment and read processing protocols as described earlier^23,24^. Variants were called using the Genome Analysis Toolkit (GATK^25,26^). Local realignment around indels and recalibration of base qualities in each sample BAM were performed with GATK version 3.4. Each sample was called with HaplotypeCaller to create gVCF files containing every position of the exome with likelihoods for variants alleles or the genomic reference allele. All variants were annotated using the Variant Quality Score Recalibration (VQSR) tool in GATK version 3.6 resulting in a VCF with germline SNVs and indels for all samples used in the analyses. The variant joint calling corresponded to the pipeline used to create the GnomAD database^23^.

### In silico analysis of SLCO1B1 variants in SUPER-Finland study

The variants of chromosome 12 were converted to Hail^27^ matrix table file format and annotated with Combined Annotation-Dependent Depletion^18,19^ (CADD, version 1.4/1.6) scores and with the Variant Effect Predictor^20^ (VEP, version 95) tool through Hail (version 0.2, reference genome GRCh38). The variants were filtered with Hail to include only those having a call rate >0.99, a HWE p-value >1e-10, an allele count >1, a genotype quality >20 and a depth >10. The *SLCO1B1* variants with a PHRED-scaled CADD score >10, predicted as missense variants by VEP and not included in the current CPIC guidelines for *SLCO1B1* were selected for further *in vitro* expression and functional analyses.

### Preparation of plasmids carrying SLCO1B1 variants

Plasmids carrying the *SLCO1B1* variants were created as described by Kiander et al.^14^. The reference *SLCO1B1* gene used was Genebank™ accession number AJ132573.1 and the mutagenesis primers are described in Supplementary Table 1. GATC Biotech’s (Constance, Germany) sequencing service confirmed the presence of the SNVs in the plasmids. Baculoviruses carrying the reference *SLCO1B1* gene, the variant *SLCO1B1* genes and the previously cloned gene for enhanced yellow fluorescent protein (eYFP) as a negative control were produced as described earlier by Tikkanen et al^28^.

### Cell culture and protein expression

HEK293 human kidney cells were cultured in Dulbecco’s Modified Eagle Medium (DMEM) and high-glucose, GlutaMax culture medium supplemented with 10% fetal bovine serum (FBS) at 37 °C, 5% CO_2_. The cells (0.5 × 10^6^) were seeded in each well of 48-well plates (ThermoFisher Scientific™ Nunc™ coated with poly-D-lysine in-house) 24 hours prior to transduction with the baculoviruses. To stimulate the expression of proteins, sodium butyrate was added with the viruses at a final concentration of 5 mM (as per in-house optimization).

### Cellular uptake assays

The cellular uptake assay was performed 48 hours post-transduction on a heated (37 °C) orbital shaker plate. For a 3-minute preincubation, transport buffer (500 µl of HBSS with 4.17 mM NaHCO3 and 25 mM HEPES adjusted to pH 7.4 with NaOH) replaced the medium in the wells. After the buffer was removed, cellular uptake began when 125 µl of 1 µM 2,7-dichlorofluorescein (DCF) in the transport buffer was added into the wells. The uptake was stopped after 15 minutes by aspiration of the test solution. Three-time wash with 500 µl ice-cold transport buffer followed and the cells were lysed with 125 µl 0.1 M NaOH. Fluorescence measurement (excitation 500 nm, emission 528 nm, bandwidth 5 nm) with the multimode microplate reader Varioskan LUX (Thermo Fisher Scientific, Vantaa, Finland) of the cell lysates was performed to quantify the DCF. 10 µl of cell lysate was mixed with 300 µl Coomassie Plus reagent and used for total protein amount quantification with absorbance analysis (595 nm) on Varioskan LUX.

### Crude membrane extraction

Baculoviruses carrying either reference or variant *SLCO1B1* and sodium butyrate (5 mM final concentration) were added to the HEK293 cells after the cells were cultured for 24 hours in T175 flasks. The cells were collected and centrifuged (3000 g, 15 min) after 48 hours and broken down with Dounce tissue homogenizer, resuspended in Tris-sucrose (TS) buffer (10 mM Tris-HEPES, 250 mM sucrose, pH 7.4) and kept on ice. After a 30-minute centrifugation the supernatant was separated and centrifuged (21,000 g, 4 °C, 99 min) again resulting in a pellet containing the crude cell membrane. The protein sample was suspended in TS buffer and quantified as previously described.

### LC–MS/MS based quantitative targeted absolute proteomics (QTAP) analysis

The LC–MS/MS-based QTAP approach was used to quantify the absolute protein expression of OATP1B1 in the crude membrane preparations. The method for the protein sample preparation and quantitation with targeted LC–MS is described earlier^14,29,30^. OATP1B1 and Na^+^/K^+^ATPase signature peptides were quantified from 50 µg of crude membrane fractions. After denaturation and break-down of the tertiary structure of the proteins, the crude membrane preparations were digested first with 1/100 LysC endopeptidase and after that with 1/100 TPCK-treated trypsin. Previously used isotope-labeled peptide mixture (3 fmol/µg protein) served as an internal standard^14^. Quantification was conducted with 6495 QQQ MS with a 1290 HPLC system and AdvanceBio peptide Map Column, 2.7 µm, 2.1 × 250 mm (Agilent Technologies, Santa Clara, CA, USA) as described previously^14,30^. The SNVs did not alter the amino acids in the analyzed peptide sequences. The peak area ratios of the analyte peptides and their respective internal standards were compared with the Skyline application (MacCoss Lab Software, Seattle, WA). The results of OATP1B1 expression are presented as relative to Na^+^/K^+^-ATPase expression level and normalized to the reference OATP1B1 protein. Absolute amount of OATP1B1 in proteomics samples is presented in Supplementary Table 2 and Supplementary Figure 1.

### Data analysis

The uptake of the DCF was normalized to the total protein amount. Uptake into eYFP-expressing cells, representing passive influx, was subtracted from uptake into OATP1B1 expressing cells, yielding active OATP1B1 mediated transport. The transport activity in OATP1B1 variant cells was then normalized to the cells expressing wild-type OATP1B1. The statistical significance of the differences in activity and expression levels was determined with one-way analysis of variance (ANOVA) with the Dunnett’s post hoc test for multiple comparisons (GraphPad Prism 6.05, GraphPad Software, San Diego, CA, USA).

### Ethics

The Coordinating Ethics Committee of the The Hospital District of Helsinki and Uusimaa (HUS) gave a favorable ethics statement (202/13/03/00/15) for the SUPER-Finland study. Prior to inclusion, written informed consent was obtained and archived from each participant. Individual-level data was pseudonymized.

### Data and code availability

The computer code used in the analysis of this study is available from the corresponding author on reasonable request and the SUPER-Finland study data is available from THL Biobank when released from the original study.

## Results

We evaluated the functionality of six SNVs of OATP1B1. The SNVs with their genomic positions, amino acid changes and locations in OATP1B1 are shown in Table 1.

**Table 1.**
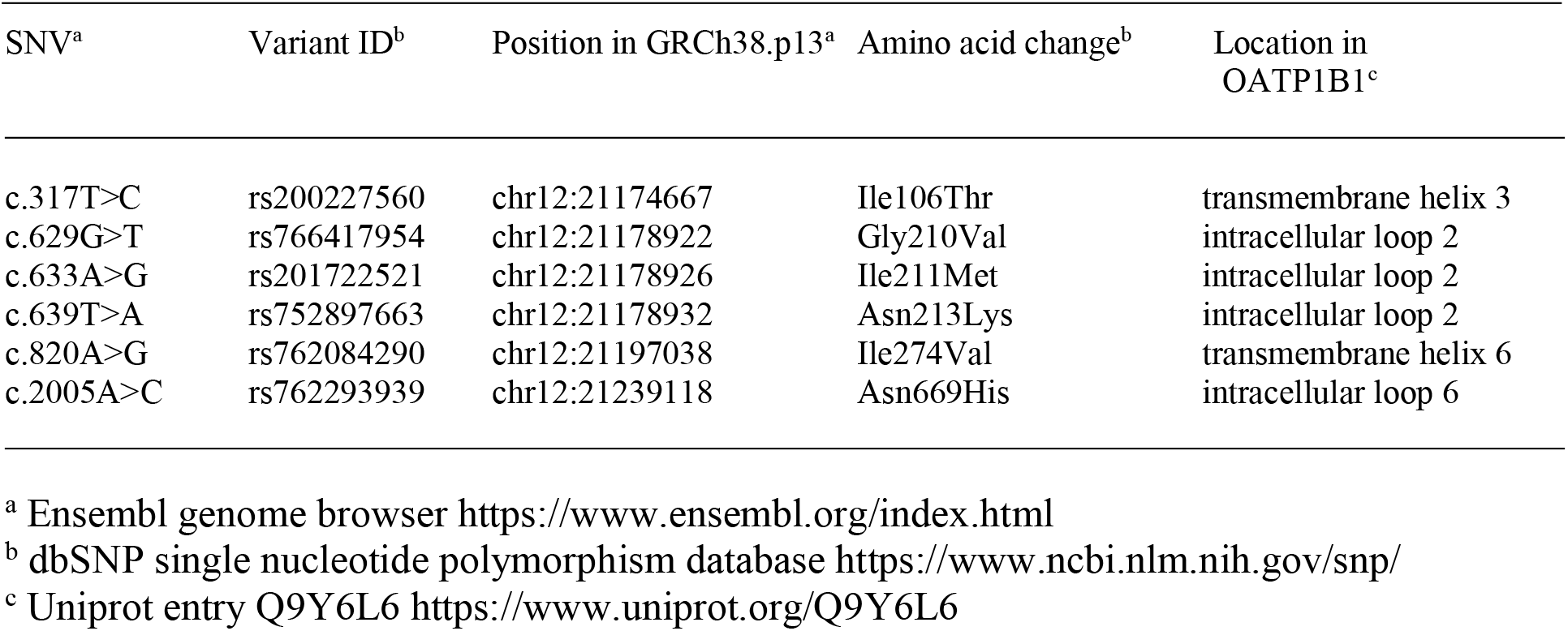
Genomic positions, amino acid changes and locations of the *SLCO1B1* SNVs.

| SNV<sup>a</sup> | Variant ID<sup>b</sup> | Position in GRCh38.p13<sup>a</sup> | Amino acid change<sup>b</sup> | Location in OATP1B1<sup>c</sup> |
| --- | --- | --- | --- | --- |
| c.317T>C | rs200227560 | chr12:21174667 | Ile106Thr | transmembrane helix 3 |
| c.629G>T | rs766417954 | chr12:21178922 | Gly210Val | intracellular loop 2 |
| c.633A>G | rs201722521 | chr12:21178926 | Ile211Met | intracellular loop 2 |
| c.639T>A | rs752897663 | chr12:21178932 | Asn213Lys | intracellular loop 2 |
| c.820A>G | rs762084290 | chr12:21197038 | Ile274Val | transmembrane helix 6 |
| c.2005A>C | rs762293939 | chr12:21239118 | Asn669His | intracellular loop 6 |

<sup>a</sup> Ensembl genome browser https://www.ensembl.org/index.html
<sup>b</sup> dbSNP single nucleotide polymorphism database https://www.ncbi.nlm.nih.gov/snp/
<sup>c</sup> Uniprot entry Q9Y6L6 https://www.uniprot.org/Q9Y6L6

The variants were expressed in HEK293 cells and the effect of the SNVs on transport activity were evaluated with a cellular uptake study using DCF as a substrate. An LC-MS/MS-based QTAP approach quantified any changes in protein abundance of OATP1B1 in crude membrane fractions.

### In silico predictions of SLCO1B1 variants in SUPER-Finland study

*In silico* predicted consequences of the six *SLCO1B1* variants are presented in Table 2 and allele frequencies of the variants are shown in Table 3.

**Table 2.**
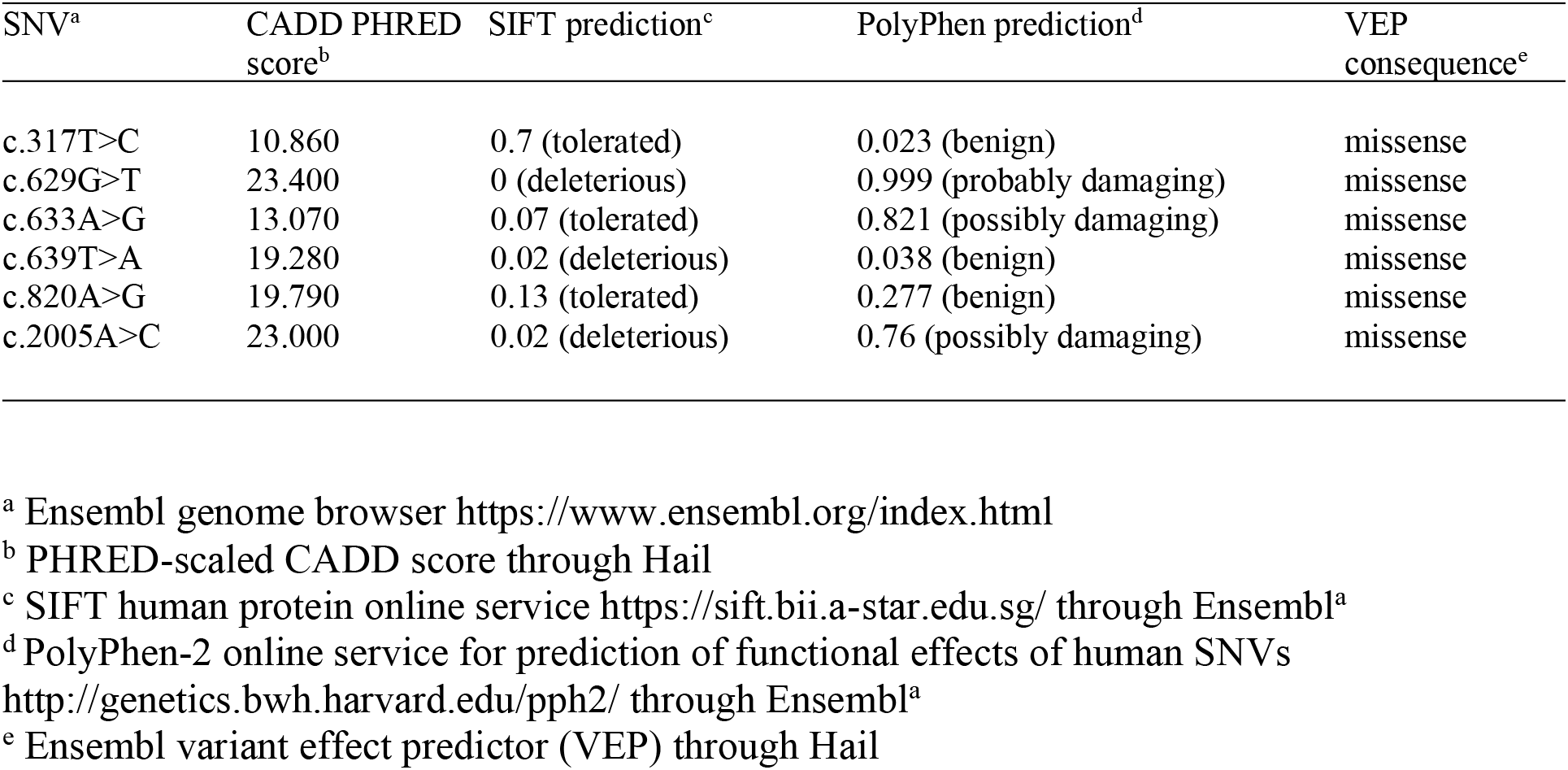
*In silico* prediction of consequences of the *SLCO1B1* SNVs.

| SNV<sup>a</sup> | CADD PHRED score<sup>b</sup> | SIFT prediction<sup>c</sup> | PolyPhen prediction<sup>d</sup> | VEP consequence<sup>e</sup> |
| --- | --- | --- | --- | --- |
| c.317T>C | 10.860 | 0.7 (tolerated) | 0.023 (benign) | missense |
| c.629G>T | 23.400 | 0 (deleterious) | 0.999 (probably damaging) | missense |
| c.633A>G | 13.070 | 0.07 (tolerated) | 0.821 (possibly damaging) | missense |
| c.639T>A | 19.280 | 0.02 (deleterious) | 0.038 (benign) | missense |
| c.820A>G | 19.790 | 0.13 (tolerated) | 0.277 (benign) | missense |
| c.2005A>C | 23.000 | 0.02 (deleterious) | 0.76 (possibly damaging) | missense |

<sup>a</sup> Ensembl genome browser https://www.ensembl.org/index.html
<sup>b</sup> PHRED-scaled CADD score through Hail
<sup>c</sup> SIFT human protein online service https://sift.bii.a-star.edu.sg/ through Ensembl<sup>a</sup>
<sup>d</sup> PolyPhen-2 online service for prediction of functional effects of human SNVs http://genetics.bwh.harvard.edu/pph2/ through Ensembl<sup>a</sup>
<sup>e</sup> Ensembl variant effect predictor (VEP) through Hail

**Table 3.** Prevalence of the *SLCO1B1* SNVs.

| SNV<sup>a</sup> | Allele Count in SUPER | Number of carriers in SUPER | Frequency in SUPER | Finnish frequency<sup>b</sup> | Non-Finnish European frequency<sup>b</sup> | Global frequency<sup>b</sup> |
| --- | --- | --- | --- | --- | --- | --- |
| c.317T>C | 4 | 4 | $2.1 \times 10^{-4}$ | $4.6 \times 10^{-5}$ | $7.4 \times 10^{-4}$ | $7.0 \times 10^{-4}$ |
| c.629G>T | 2 | 2 | $1.1 \times 10^{-4}$ | $2.3 \times 10^{-4}$ | 0.0 | $3.2 \times 10^{-5}$ |
| c.633A>G | 2 | 2 | $1.1 \times 10^{-4}$ | 0.0 | $8.8 \times 10^{-6}$ | $3.6 \times 10^{-3}$ |
| c.639T>A | 16 | 16 | $8.5 \times 10^{-4}$ | $8.9 \times 10^{-4}$ | 0.0 | $7.6 \times 10^{-5}$ |
| c.820A>G | 2 | 2 | $1.1 \times 10^{-4}$ | $4.6 \times 10^{-5}$ | 0.0 | $4.0 \times 10^{-6}$ |
| c.2005A>C | 2 | 2 | $1.1 \times 10^{-4}$ | $2.3 \times 10^{-4}$ | 0.0 | $2.0 \times 10^{-5}$ |

<sup>a</sup> Ensembl genome browser https://www.ensembl.org/index.html
<sup>b</sup> Exome population frequencies are retrieved from the gnomAD browser version 2.1.1 https://gnomad.broadinstitute.org/
Allele frequencies are counted by dividing allele count by overall number of alleles in a population.

SUPER-Finland subjects are heterozygous for the studied variants. The most frequent SNVs were c.639T>A, which was found in 16 subjects and c.317T>C, which was found in four subjects. The other four SNVs were found in only two subjects. Based on the *in silico* predictions by CADD, SIFT and PolyPhen, c.629G>T and c.2005A>C were most likely to have a damaging effect, and c.317T>C to be tolerated, while the predictions on the three other variants were more conflicting. All the variants are predicted to be missense variants by VEP. Two-dimensional prediction of the OATP1B1 transporter with the amino acid locations of the identified variants is presented in Figure 1.

**Figure 1.**
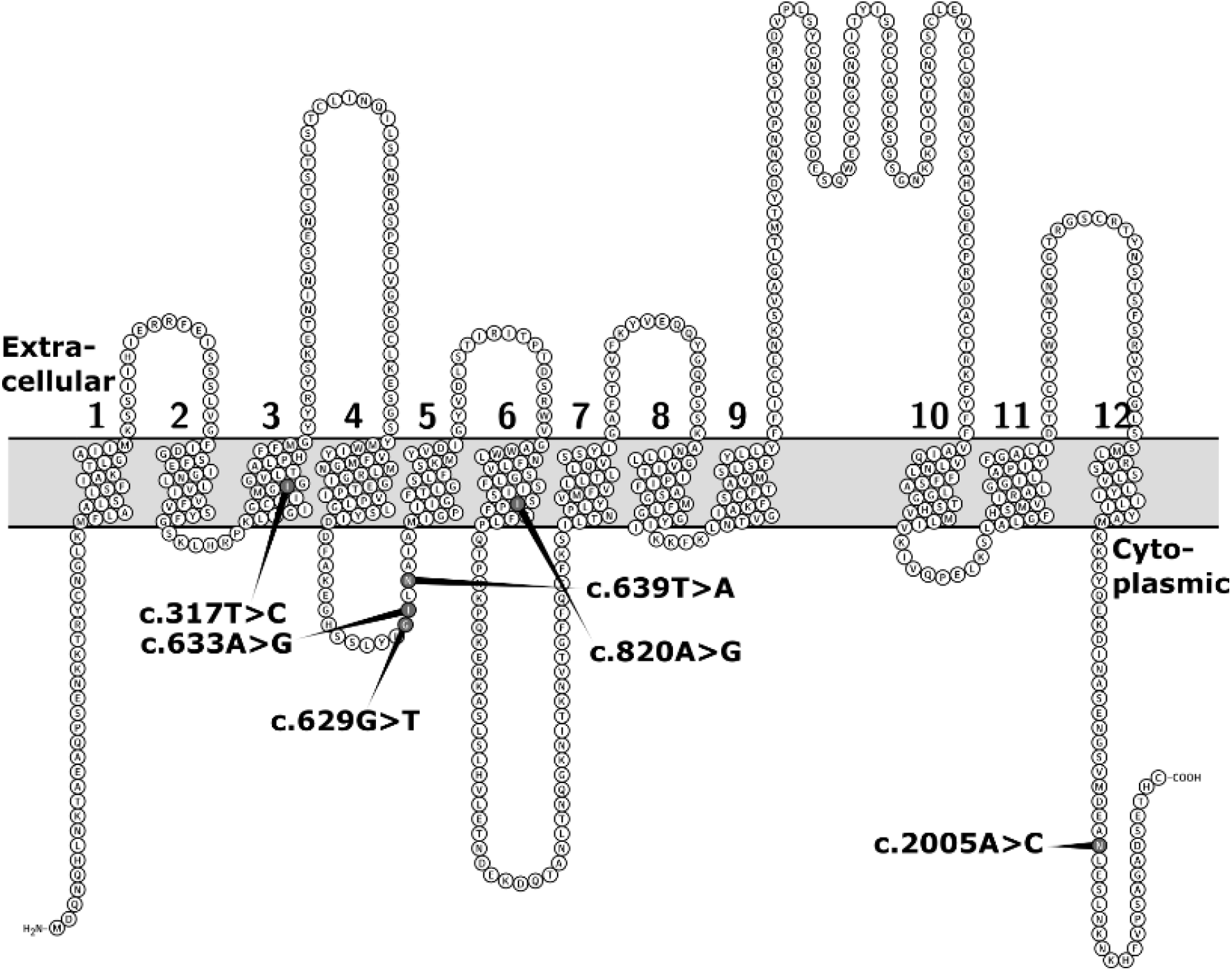
Location of the amino acids affected by the identified *SLCO1B1* SNVs in two-dimensional prediction of OATP1B1 transporter. Numbers 1-12 denote the putative transmembrane helices. The figure is based on the Uniprot entry Q9Y6L6 and generated with Protter^31^.

### In vitro transport activity and membrane protein expression

We discovered that c.629G>T (p.G210V) abolishes the transport activity of OATP1B1 (Figure 2). Membrane protein abundance was also significantly decreased by approximately 70% compared to the OATP1B1 reference. The other variants did not alter transport activity or membrane protein expression to a statistically significant degree even though the activity was on average higher than the reference (Figure 2).

**Figure 2.**
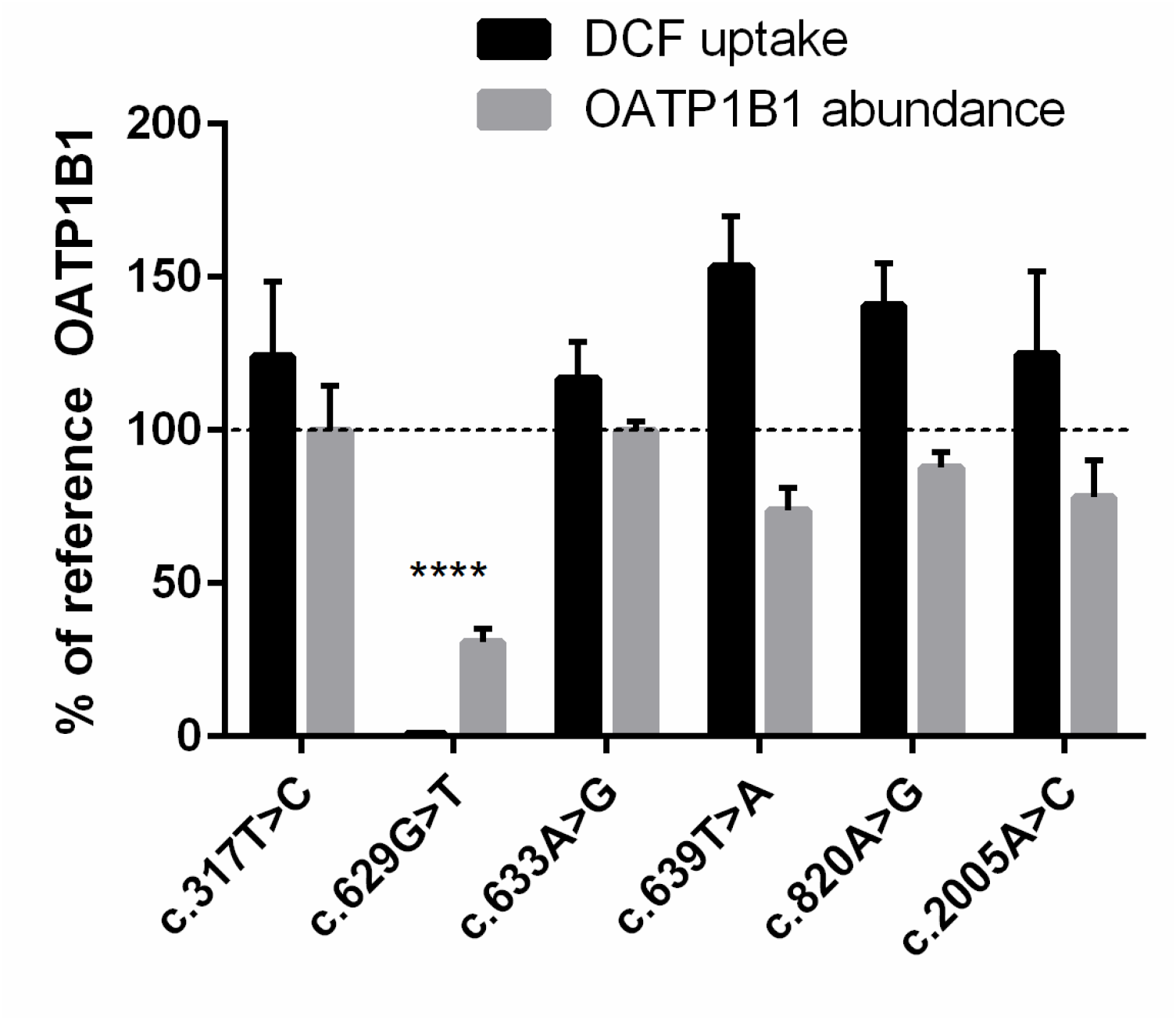
OATP1B1 mediated transport of 1 µM DCF into HEK293 cells over 15 minutes: Results are calculated as mean of four experiments conducted in quadruplicates and represented % of reference OATP1B1 transport ±SEM (n=4). LC-MS/MS proteomics analysis of 50 µg HEK293 crude membrane preparations expressing variant OATP1B1: Abundance of OATP1B1 was quantified in four independent samples, normalized to Na+/K+-ATPase and reference OATP1B1 abundance (100%). The results are presented as mean ±SEM (n=4). **** = P<0.0001 (compared to the reference) ANOVA + Dunnett’s post hoc test.

Interestingly, though, c.639T>A (p.213N>K) seemed to enhance transport activity, which was on average 162% of reference OATP1B1 transport but failed to reach statistical significance. Protein expression of c.639T>A (p.213N>K), however, was not increased.

## Discussion

We have provided novel functional annotations for six *SLCO1B1* variants. Among these, c.629G>T (p.210G>V) was found to be a loss-of-function variant. The variant is in the intracellular loop of the OATP1B1 transporter, and according to GnomAD^23^ the variant has been detected only in Finnish and South Asian populations. We used computational algorithms to prioritize *SLCO1B1* variants for *in vitro* validation. The PHRED-scaled CADD score for this variant was 23.4 and it was predicted to be deleterious by SIFT and probably damaging by PolyPhen. A variant with a similar CADD score of 23.0 (c.2005A>C, p.N669H) and classified as deleterious and possibly damaging by SIFT and PolyPhen, respectively, had normal function according to our *in vitro* studies. This underlines the difficulty of computational functional annotation of missense variants. Due to the enormous number of variants in whole exome sequencing studies, these studies need to rely on computational functional predictions, as not every potential loss-of-function variant throughout the genome can be tested in the laboratory. The definition of a computationally-predicted loss-of-function variant is a matter of sensitivity and specificity. Selecting a conservative definition would filter out potential disease-causing mutations, while too liberal a threshold creates noise in association signals. As CADD score is based on regional mutational constraint^18,19^, it might not be optimal in capturing pharmacogenetic variants as genes responsible for metabolism and transport of xenobiotic compounds might not be under as strong selection as genes related to, for example, brain development.

*SLCO1B1* c.629G>T results in the replacement of a glycine with a valine in position 210, located at the end of the predicted intracellular loop 2 (Figure 1). Glycine has no side-chain and has a very high tendency to build turns in the secondary structure of polypeptide chains^32^. Valine, on the other hand, has a preference to form strands, and obstructs the formation of turns and bends in the secondary structure. Thus, it is understandable that this substitution can impair the proper folding and membrane localization of OATP1B1.

*SLCO1B1* c.317T>C (p.106I>T), c.633A>G (p.211I>M) and c.820A>G (p.274I>V) are all in codons that originally code for isoleucine but the SNVs result in substitutions with amino acids with similar properties. Similar to isoleucine, valine and threonine have a preference for forming strands in the secondary structure of proteins. Additionally, methionine is neutral to strand formation and thus these variants most likely do not alter the secondary structure to a significant degree.

*SLCO1B1* c.639T>A (p.213N>K) changes an amidic asparagine into a basic lysine, which in physiological pH has a positive charge. While the actual three-dimensional structure of the OATP proteins remain unknown, OATPs are predicted to form a positively charged pore^33^. Since many OATP1B1 substrates are anionic compounds, an additional positive charge near this putative pore in the intracellular loop where c.639T>A (p.N213K) is located might increase substrate affinity. According to GnomAD^23^ the c.639T>A variant has been identified only in Finnish population. A similar gain-of-charge occurs in *SLCO1B1* c.1007C>G (p.P336R, located in TMH 7) and was shown *in vitro* to have substrate-dependent normal to enhanced OATP1B1 transport activity^14,34^. Likewise, *SLCO1B1* c.2005A>C (p.669N>H) also gains a charge and increases activity slightly without increasing protein expression (Figure 2). All things considered, additional *in vitro* studies with multiple substrates and concentration-dependency assays are required to acquire supplementary mechanistic information on the effect of c.639T>A (p.213N>K) on OATP1B1 activity and its possible gain-of-function.

An increased-function phenotype has been reported for the *SLCO1B1*: *\*14* haplotype (that includes SNVs c.388A>G and 463C>A) increases OATP1B1 protein expression, decreases the exposure to atorvastatin and simvastatin acid, and enhances the LDL-lowering effect of fluvastatin^35,36,37,38^. Therefore, it is possible that the c.639T>A (p.213N>K) SNV characterized in this study might similarly increase hepatic intrinsic influx clearance of at least some substrate drugs *in vivo*, but this remains to be clarified. In the case of statins, where liver is the target organ, increased hepatic uptake and decreased systemic exposure are unlikely to have a negative impact. However, for substrates that are dependent on adequate systemic concentrations for their pharmacological action, such as methotrexate, a gain-of-function phenotype of OATP1B1 could reduce their efficacy and possibly require higher doses to reach the desired clinical outcome^39^.

Polypharmacy is common in patients treated with antipsychotic medication, leading to a high probability of adverse drug affects and drug-drug interactions^40^. While to date, according to The University of Washington Drug Interaction Database (DIDB)^41^, no central nervous system agents have been identified as OATP1B1 substrates, many other drugs can be prescribed to these patients to treat their somatic diseases. Among these are often statins to treat dyslipidemias which are associated with antipsychotic medications^9^. The association between statin-use and muscle symptoms has been thoroughly investigated. Variants in *SLCO1B1* are associated with myopathy in particular when there is a documented increase in creatinine kinase concentration in blood^3,42^. A poor function phenotype of OATP1B1 can increase the probability for adverse effects during statin treatment thus reducing medication adherence.

The *SLCO1B1*5* and *SLCO1B1*15* haplotypes that contain the function-impairing c.521T>C SNV are associated with reduced hepatic clearance of substrate drugs, increased systemic exposure and increased risk of SAMS^1,6,7^. The current gene-based prescribing guideline from the Clinical Pharmacogenetics Implementation Consortium (CPIC) states that individuals with decreased and poor function phenotypes of OATP1B1 should limit statin doses and those with poor function phenotypes should avoid simvastatin altogether^6^. Based on our *in vitro* studies, c.629G>T (p.G210G>V) could be categorized as a poor function phenotype, since the *in vitro* activity and abundance is comparable to the *SLCO1B1* c.521T>C genotype^1,14^. However, homozygous carriers of a rare variant like c.629G>T (p.G210G>V) are very uncommon. It is more likely to occur in a heterozygous state as observed in our study population (Table 3), or, like c.521T>C in *SLCO1B1*15*, in combination with c.388A>G, which has a frequency of approximately 26-77% depending on the population^43^. Additionally, c.521T>C allele is quite common in certain populations, for example the minor allele frequency in Finnish Europeans is 0.21^23^. Consequently, even as many as every fourth c.629G>T carrier is likely to carry c.521T>C resulting in a poor function phenotype. While the heterozygous genotype would limit the clinical consequences from a poor function phenotype to a decreased function phenotype, these individuals would still have an increased risk of SAMS during high-dose statin treatment and thus could benefit from genotype-guided dosing.

## Conclusions

We have identified a novel *SLCO1B1* loss-of-function mutation which could be accounted for in guidelines describing individualized lipid-lowering therapy. The variant c.629G>T (p.210G>V) abolishes the transport activity of OATP1B1 and may predispose patients to increased risk of SAMS during statin treatment. This emphasizes the need to include diverse populations in drug trials as adverse effects related to population-specific variants might otherwise be missed. The other five functionally-characterized *SLCO1B1* variants (c.317T>C, c.633A>G, c.639T>A, c.820A>G and c.2005A>C) can be considered normal-function variants. Protein expression was not altered but DCF uptake was clearly enhanced, although failed to reach statistical significance, by the variant c.639T>A (p.213N>L). To study the possible gain-of-function effect on OATP1B1 activity of this variant, further *in vitro* studies with multiple substrates and concentration dependency assays are required.

## Supporting information

STROBE_checklist

## Acknowledgements

We thank the SUPER-Finland study participants. We would like to thank the collaborating SUPER-Finland study group: Mark Daly, Steve Hyman, Amanda Elliott, Benjamin Neale, Willehard Haaki, Teemu Männynsalo, Tuomas Jukuri, Kimmo Suokas, Jussi Niemi-Pynttäri, Asko Wegelius, Risto Kajanne, Aija Kyttälä, Noora Ristiluoma, Hannu Turunen, Auli Toivola, Annamari Tuulio-Henriksson, Tiina Paunio, Juha Veijola, Katriina Hakakari, Elina Hietala, Jaakko Keinänen, Martta Kerkelä, Tuomo Kiiskinen, Sari Lång-Tonteri, Nina Lindberg, Ville Mäkipelto, Atiqul Mazumder, Erik Cederlöf, Zuzanna Misiewicz, Julia Moghadampour, Arto Mustonen, Solja Niemelä, Olli Pietiläinen, Susanna Rask, Elmo Saarentaus, Anssi Solismaa, Andre Sourander, Marjo Taivalantti and Imre Västrik for the SUPER study data and support. We would like to thank Marju Monnonen, Natalija Trunkelj, Noora Sjöstedt and Laura Suominen for technical assistance as well as Dr. Yasuo Uchida for consultation on sample preparation of the proteomics samples. Dr. Mikko Gynther is acknowledged for the methodology in proteomics. We acknowledge the Drug Discovery and Chemical Biology Network for providing access to screening instrumentation and the Genomics Platform of the Broad Institute of MIT and Harvard for whole exome sequencing the SUPER-Finland study samples.

## Author contribution

Conceptualization: M.N., A.V.A.; Data curation: K.H., W.K., K-S.V., L.U., J.L., SUPER-Finland researchers, A.V.A.; Formal analysis: K.H., W.K., H.K., M.N., A.V.A.; Funding acquisition: K.H., M.L., J.T., A.P., S.A., M.N., A.V.A.; Investigation: K.H., W.K., SUPER-Finland researchers, A.V.A.; Methodology: H.K., M.N., A.P., S.A., A.V.A.; Project administration : A.P., M.N., A.V.A.; Resources: H.K., M.N., J.T., S.A., A.P.; Software: K.H., L.U., J.L., A.V.A.; Supervision: H.K., M.L., J.T., M.N., A.V.A.; Validation: W.K., K-S.V., S.A.; Visualization: K.H., W.K.; Writing – original draft: K.H., W.K., H.K., M.N., A.V.A.; Writing – review & editing: K.H., W.K., H.K., M.L., L.U., K.-S.V., S.A., M.H., K.L., O.K., E.I., T.K., J.L., J.S., J.H., J.T., A.P., M.N., A.V.A.

## References

1. Niemi, M., Pasanen, M. K. & Neuvonen, P. J. Organic Anion Transporting Polypeptide 1B1: a Genetically Polymorphic Transporter of Major Importance for Hepatic Drug Uptake. Pharmacol. Rev. 63, 157–181 (2011).

2. König, J., Cui, Y., Nies, A. T. & Keppler, D. A novel human organic anion transporting polypeptide localized to the basolateral hepatocyte membrane. Am. J. Physiol. Gastrointest. Liver Physiol. 278, G156–G164 (2000).

3. Meade, T. et al. SLCO1B1 variants and statin-induced myopathy - A genomewide study. N. Engl. J. Med. 359, 789–799 (2008).

4. Niemi, M. Transporter pharmacogenetics and statin toxicity. Clin. Pharmacol. Ther. 87, 130–133 (2010).

5. Ramsey, L. et al. The Clinical Pharmacogenetics Implementation Consortium Guideline for SLCO1B1 and Simvastatin-Induced Myopathy: 2014 Update. Clin. Pharmacol. Ther. 96, 423–428 (2014).

6. Cooper-Dehoff, R. M. et al. The Clinical Pharmacogenetics Implementation Consortium Guideline for SLCO1B1, ABCG2, and CYP2C9 genotypes and Statin-Associated Musculoskeletal Symptoms. Clin. Pharmacol. Ther. 111, 1007–1021 (2022).

7. Turner, R. M. & Pirmohamed, M. Statin-Related Myotoxicity: A Comprehensive Review of Pharmacokinetic, Pharmacogenomic and Muscle Components. J. Clin. Med. 9, 1–37 (2019).

8. Casey, D. E. et al. Antipsychotic-Induced Weight Gain and Metabolic Abnormalities: Implications for Increased Mortality in Patients With Schizophrenia Table 1. ‘The Metabolic Syndrome’ According to the Adult Treatment Panel III a. J Clin Psychiatry 65, 4–18 (2004).

9. Chang, S.-C., Kheng Goh, K. & Lu, M.-L. Metabolic disturbances associated with antipsychotic drug treatment in patients with schizophrenia: State-of-the-art and future perspectives. World J. Psychiatry 11, 696–710 (2021).

10. U.S. Food and Drug Administration. Clinical Drug Interaction Studies — Cytochrome P450 Enzyme and Transporter-Mediated Drug Interactions Guidance for Industry. (2020). Available at: https://www.fda.gov/regulatory-information/search-fda-guidance-documents/clinical-drug-interaction-studies-cytochrome-p450-enzyme-and-transporter-mediated-drug-interactions.

11. European Medicines Agency. Guideline on the investigation of drug interactions. (2012). Available at: https://www.ema.europa.eu/en/documents/scientific-guideline/guideline-investigation-drug-interactions-revision-1_en.pdf.

12. Lauschke, V. M. & Ingelman-Sundberg, M. Emerging strategies to bridge the gap between pharmacogenomic research and its clinical implementation. npj Genomic Medicine 5, 1–7 (2020).

13. Ramsey, L. B. et al. Rare versus common variants in pharmacogenetics: SLCO1B1 variation and methotrexate disposition. Genome Res. 22, 1–8 (2012).

14. Kiander, W. et al. The Effect of Single Nucleotide Variations in the Transmembrane Domain of OATP1B1 on in vitro Functionality. Pharm. Res. 38, 1663–1675 (2021).

15. Seaby, E. G., Pengelly, R. J. & Ennis, S. Exome sequencing explained: a practical guide to its clinical application. Brief. Funct. Genomics 5, 374–384 (2016).

16. Lauschke, V. M. & Ingelman-Sundberg, M. Precision Medicine and Rare Genetic Variants. Trends Pharmacol. Sci. 37, 85–86 (2016).

17. Kozyra, M., Ingelman-Sundberg, M. & Lauschke, V. M. Rare genetic variants in cellular transporters, metabolic enzymes, and nuclear receptors can be important determinants of interindividual differences in drug response. Genet. Med. 19, 20–29 (2017).

18. Kircher, M. et al. A general framework for estimating the relative pathogenicity of human genetic variants. Nat. Genet. 46, 310–315 (2014).

19. Rentzsch, P., Witten, D., Cooper, G. M., Shendure, J. & Kircher, M. CADD: predicting the deleteriousness of variants throughout the human genome. Nucleic Acids Res. 47, D886–D894 (2019).

20. Mclaren, W. et al. The Ensembl Variant Effect Predictor. Genome Biol. 17, 2–14 (2016).

21. Hovatta I., Terwilliger J., Lichtermann D., Mäkikyrö T., Suvisaari J., Peltonen L., L. Schizophrenia in the genetic isolate of Finland. American Journal of Medical Genetics 353–360 (1997).

22. Li, H. & Durbin, R. Fast and accurate short read alignment with Burrows-Wheeler transform. Bioinformatics 25, 1754–1760 (2009).

23. Karczewski, K. J. et al. The mutational constraint spectrum quantified from variation in 141,456 humans. Nature 581, 434–443 (2020).

24. Lek, M. et al. Analysis of protein-coding genetic variation in 60,706 humans. Nature 536, 285–291 (2016).

25. Van Der Auwera, G. A. et al. From FastQ data to high confidence variant calls: the Genome Analysis Toolkit best practices pipeline. Curr Protoc Bioinforma. 11(1110), 11.10.1–11.10.33 (2013).

26. Mckenna, A. et al. The Genome Analysis Toolkit: A MapReduce framework for analyzing next-generation DNA sequencing data. Genome Res. 20, 1297–1303 (2010).

27. Hail Team. Hail 0.2. https://github.com/hail-is/hail

28. Tikkanen, A. et al. Food Additives as Inhibitors of Intestinal Drug Transporter OATP2B1. Mol. Pharm. 17, 3748–3758 (2020).

29. Uchida, Y. et al. A study protocol for quantitative targeted absolute proteomics (QTAP) by LC-MS/MS: application for inter-strain differences in protein expression levels of transporters, receptors, claudin-5, and marker proteins at the blood-brain barrier in ddY, FVB, and. Fluids Barriers CNS 10, 1–22 (2013).

30. Huttunen, J., Gynther, M., Vellonen, K.-S. & Huttunen, K. M. L-Type amino acid transporter 1 (LAT1)-utilizing prodrugs are carrier-selective despite having low affinity for organic anion transporting polypeptides (OATPs). Int. J. Pharm. 571, 118714 (2019).

31. Omasits, U., Ahrens, C. H., Müller, S. & Wollscheid, B. Protter: interactive protein feature visualization and integration with experimental proteomic data. Bioinformatics 30, 884–886 (2014).

32. Malkov, S. N., Živković, M. V., Beljanski, M. V., Stojanović, S. D. & Zarić, S. D. A reexamination of correlations of amino acids with particular secondary structures. Protein J. 28, 74–86 (2009).

33. Meier-Abt, F., Mokrab, Y. & Mizuguchi, K. Organic anion transporting polypeptides of the OATP/SLCO superfamily: Identification of new members in nonmammalian species, comparative modeling and a potential transport mode. J. Membr. Biol. 208, 213–227 (2006).

34. Kameyama, Y., Yamashita, K., Kobayashi, K., Hosokawa, M. & Chiba, K. Functional characterization of SLCO1B1 (OATP-C) variants, SLCO1B1*5, SLCO1B1*15 and SLCO1B1*15 + C1007G, by using transient expression systems of HeLa and HEK293 cells. Pharmacogenet. Genomics 15, 513–522 (2005).

35. Couvert, P. et al. Association between a frequent allele of the gene encoding OATP1B1 and enhanced LDL-lowering response to fluvastatin therapy. Pharmacogenomics 9, 1217–1227 (2008).

36. Nies, A. T. et al. Genetics is a major determinant of expression of the human hepatic uptake transporter OATP1B1, but not of OATP1B3 and OATP2B1. Genome Med. 5, (2013).

37. Prasad, B. et al. Interindividual Variability in Hepatic Organic Anion-Transporting Polypeptides and P-Glycoprotein (ABCB1) Protein Expression: Quantification by Liquid Chromatography Tandem Mass Spectroscopy and Influence of Genotype, Age, and Sex. Drug Metab Dispos 42, 78–88 (2014).

38. Mykkänen, A. J. H. et al. Genomewide Association Study of Simvastatin Pharmacokinetics. Clin. Pharmacol. Ther. 0, 1–11 (2022).

39. Ramsey, L. B. et al. Association of SLCO1B1 *14 Allele with Poor Response to Methotrexate in Juvenile Idiopathic Arthritis Patients. ACR Open Rheumatol. 1, 58–62 (2019).

40. Fornaro, M. et al. Prevalence and clinical features associated with bipolar disorder polypharmacy: a systematic review. Neuropsychiatr. Dis. Treat. 12, 719–735 (2016).

41. Hachad, H., Ragueneau-Majlessi, I. & Levy, R. H. A useful tool for drug interaction evaluation: The University of Washington Metabolism and Transport Drug Interaction Database. Hum. Genomics 5, 61–72 (2010).

42. Danik, J. S. et al. Lack of association between SLCO1B1 polymorphisms and clinical myalgia following rosuvastatin therapy. Am. Heart J. 165, 1008–1014 (2013).

43. Pasanen, M. K., Neuvonen, P. J. & Niemi, M. Global analysis of genetic variation in SLCO1B1. Pharmacogenomics 9, 19–33 (2008).

