## Supplementary data for "Functional characterization of six *SLCO1B1* (OATP1B1) variants observed in Finnish individuals with a psychotic disorder"

**Supplementary Table I** Mutagenesis primers used in creating SNVs in the SLCO1B1 gene. Small case letter denotes the nucleotide substitution.

| SNV | Amino acid change | Forward | Reverse |
|---|---|---|---|
| c.317T>C | Ile106Thr | ATTATGGGAAcTGGAGGTGTT | GAAACAACCGATTCCAATTAAC |
| c.629G>T | Gly210Val | ATTTAGGTATgTTGAATGCAATAG | ACAAAGAAGAATGTCCTTC |
| c.633A>G | Ile211Met | TTGTATTTAGtTATATTGAATGCAATAG | AGAAGAATGTCCTTCTTTAG |
| c.639T>A | Asn213Lys | GTATATTGAAaGCAATAGCAATGATTG | CTAAATACAAAGAAGAATGTCC |
| c.820A>G | Ile274Val | TATTTCTTCCgTACCATTCTTTTTC | ATGGAGAATAGTCCAGAC |
| c.2005A>C | Asn669His | GGATGAAGCAcACTTAGAATCC | ATGACACTTCCATTTTCTG |

**Supplementary Table II** Absolute amount of OATP1B1 in proteomics samples.
$Na^+/K^+$/ATPase unique peptide: AAVPDAVGK, OATP1B1 unique peptide: LNTVGIAK

| Variant | Average OATP1B1 abundance fmol/µg protein | Standard error of the mean | Average $Na^+/K^+$/ATPase abundance fmol/µg protein | Standard error of the mean |
|---|---|---|---|---|
| Reference | 1.09 | 0.52 | 2.45 | 0.59 |
| I106T | 1.09 | 0.49 | 2.61 | 0.58 |
| G210V | 0.21 | 0.059 | 1.81 | 0.27 |
| I211M | 0.95 | 0.27 | 2.49 | 0.51 |
| N213K | 0.63 | 0.29 | 1.84 | 0.23 |
| I274V | 0.73 | 0.29 | 1.92 | 0.14 |
| N669H | 0.62 | 0.23 | 1.92 | 0.36 |

**Supplementary Figure 1** Absolute amount of OATP1B1 and $Na^+/K^+$/ATPase in proteomics samples: individual samples coupling the abundance of two unique peptides AAVPDAVGK for $Na^+/K^+$/ATPase and LNTVGIAK for OATP1B1.

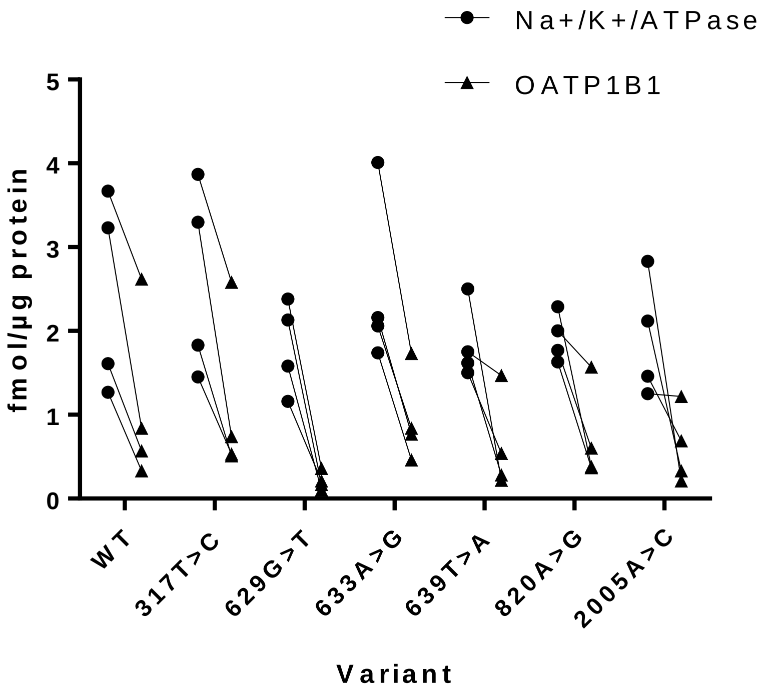
